# Global Biobank Meta-analysis Initiative: powering genetic discovery across human diseases

**DOI:** 10.1101/2021.11.19.21266436

**Authors:** Wei Zhou, Masahiro Kanai, Kuan-Han H Wu, Rasheed Humaira, Kristin Tsuo, Jibril B Hirbo, Ying Wang, Arjun Bhattacharya, Huiling Zhao, Shinichi Namba, Ida Surakka, Brooke N Wolford, Valeria Lo Faro, Esteban A Lopera-Maya, Kristi Läll, Marie-Julie Favé, Sinéad B Chapman, Juha Karjalainen, Mitja Kurki, Maasha Mutaamba, Ben M Brumpton, Sameer Chavan, Tzu-Ting Chen, Michelle Daya, Yi Ding, Yen-Chen A Feng, Christopher R Gignoux, Sarah E Graham, Whitney E Hornsby, Nathan Ingold, Ruth Johnson, Triin Laisk, Kuang Lin, Jun Lv, Iona Y Millwood, Priit Palta, Anita Pandit, Michael Preuss, Unnur Thorsteinsdottir, Jasmina Uzunovic, Matthew Zawistowski, Xue Zhong, Archie Campbell, Kristy Crooks, Geertruida h De Bock, Nicholas J Douville, Sarah Finer, Lars G Fritsche, Christopher J Griffiths, Yu Guo, Karen A Hunt, Takahiro Konuma, Riccardo E Marioni, Jansonius Nomdo, Snehal Patil, Nicholas Rafaels, Anne Richmond, Jonathan A Shortt, Peter Straub, Ran Tao, Brett Vanderwerff, Kathleen C Barnes, Marike Boezen, Zhengming Chen, Chia-Yen Chen, Judy Cho, George Davey Smith, Hilary K Finucane, Lude Franke, Eric Gamazon, Andrea Ganna, Tom R Gaunt, Tian Ge, Hailiang Huang, Jennifer Huffman, Clara Lajonchere, Matthew H Law, Liming Li, Cecilia M Lindgren, Ruth JF Loos, Stuart MacGregor, Koichi Matsuda, Catherine M Olsen, David J Porteous, Jordan A Shavit, Harold Snieder, Richard C Trembath, Judith M Vonk, David Whiteman, Stephen J Wicks, Cisca Wijmenga, John Wright, Jie Zheng, Xiang Zhou, Philip Awadalla, Michael Boehnke, Nancy J Cox, Daniel H Geschwind, Caroline Hayward, Kristian Hveem, Eimear E Kenny, Yen-Feng Lin, Reedik Mägi, Hilary C Martin, Sarah E Medland, Yukinori Okada, Aarno V Palotie, Bogdan Pasaniuc, Serena Sanna, Jordan W Smoller, Kari Stefansson, David A van Heel, Robin G Walters, Sebastian Zoellner, Biobank Japan, BioMe, BioVU, Canadian Partnership for Tomorrow, China Kadoorie Biobank Collaborative Group, Colorado Center for Personalized Medicine, deCODE Genetics, Estonian Biobank, FinnGen, Generation Scotland, Genes & Health, LifeLines, Mass General Brigham Biobank, Michigan Genomics Initiative, QIMR Berghofer Biobank, Taiwan Biobank, The HUNT Study, UCLA ATLAS Community Health Initiative, UK Biobank, Alicia R Martin, Cristen J Willer, Mark J Daly, Benjamin M Neale

**Author notes:** These authors jointly supervised this work (Cristen J Willer, Mark J Daly, Benjamin M Neale).

## Abstract

Biobanks are being established across the world to understand the genetic, environmental, and epidemiological basis of human diseases with the goal of better prevention and treatments. Genome-wide association studies (GWAS) have been very successful at mapping genomic loci for a wide range of human diseases and traits, but in general, lack appropriate representation of diverse ancestries - with most biobanks and preceding GWAS studies composed of individuals of European ancestries. Here, we introduce the Global Biobank Meta-analysis Initiative (GBMI) -- a collaborative network of 19 biobanks from 4 continents representing more than 2.1 million consented individuals with genetic data linked to electronic health records. GBMI meta-analyzes summary statistics from GWAS generated using harmonized genotypes and phenotypes from member biobanks. GBMI brings together results from GWAS analysis across 6 main ancestry groups: approximately 33,000 of African ancestry either from Africa or from admixed-ancestry diaspora (AFR), 18,000 admixed American (AMR), 31,000 Central and South Asian (CSA), 341,000 East Asian (EAS), 1.4 million European (EUR), and 1,600 Middle Eastern (MID) individuals. In this flagship project, we generated GWASs from across 14 exemplar diseases and endpoints, including both common and less prevalent diseases that were previously understudied. Using the genetic association results, we validate that GWASs conducted in biobanks worldwide can be successfully integrated despite heterogeneity in case definitions, recruitment strategies, and baseline characteristics between biobanks. We demonstrate the value of this collaborative effort to improve GWAS power for diseases, increase representation, benefit understudied diseases, and improve risk prediction while also enabling the nomination of disease genes and drug candidates by incorporating gene and protein expression data and providing insight into the underlying biology of the studied traits.

## Introduction

Understanding the genetic basis of disease can elucidate the biology or underlying epidemiological risk factors, nominate genes as drug targets, and identify at-risk individuals for prevention strategies. Genetic association studies have been routinely performed genome-wide for over 15 years and have identified thousands of loci for hundreds of diseases and traits (see GWAS Catalog (MacArthur et al., 2017)). Meta-analysis across cohorts has been instrumental in making these discoveries. However, most genomics research has been performed primarily in cohorts of European ancestries and conducted in mostly high-resource countries. Although much remains to be done to address the lack of representation in genomics, here we present the Global Biobank Meta-analysis Initiative (GBMI), a small step in building a more comprehensive view of the impact of genetic variation on human health and disease.

Biobanks with health data linked with genomic information provide unprecedented resources for the genetic research community. The rapid drop in the cost of genotyping and sequencing has led to an increase in the number of genomically profiled biobanks worldwide. Compared to disease or trait-based cohorts centered around a particular phenotype or several relevant phenotypes, biobanks enable cost-effective genetic discovery for hundreds to thousands of phenotypes, curated from electronic health records (EHRs), registry-based data (e.g. pharmaceutical, death, or cancer registry data), and/or epidemiological questionnaires to understand the genetic etiology of human diseases (Bowton et al., 2014; Wolford et al., 2018).

In 2019, we formed the GBMI bringing together 19 biobanks to work together to understand the genetic basis of human health and disease (**Figure 1, Supplementary Table 1**). The goal was to jumpstart and align global efforts, particularly since meta-analysis of GWAS is simple in terms of data sharing yet enables a variety of scientific goals including: increasing the power of GWASs for common diseases, enabling the genetic investigation into less prevalent or understudied diseases, increasing the ancestral diversity of genetic association studies and in doing so analyzing a broader set of genetic variation, cross-validating new findings across biobanks, and facilitating follow-up analyses such as polygenic risk scores or Mendelian Randomization.

**Figure 1.**
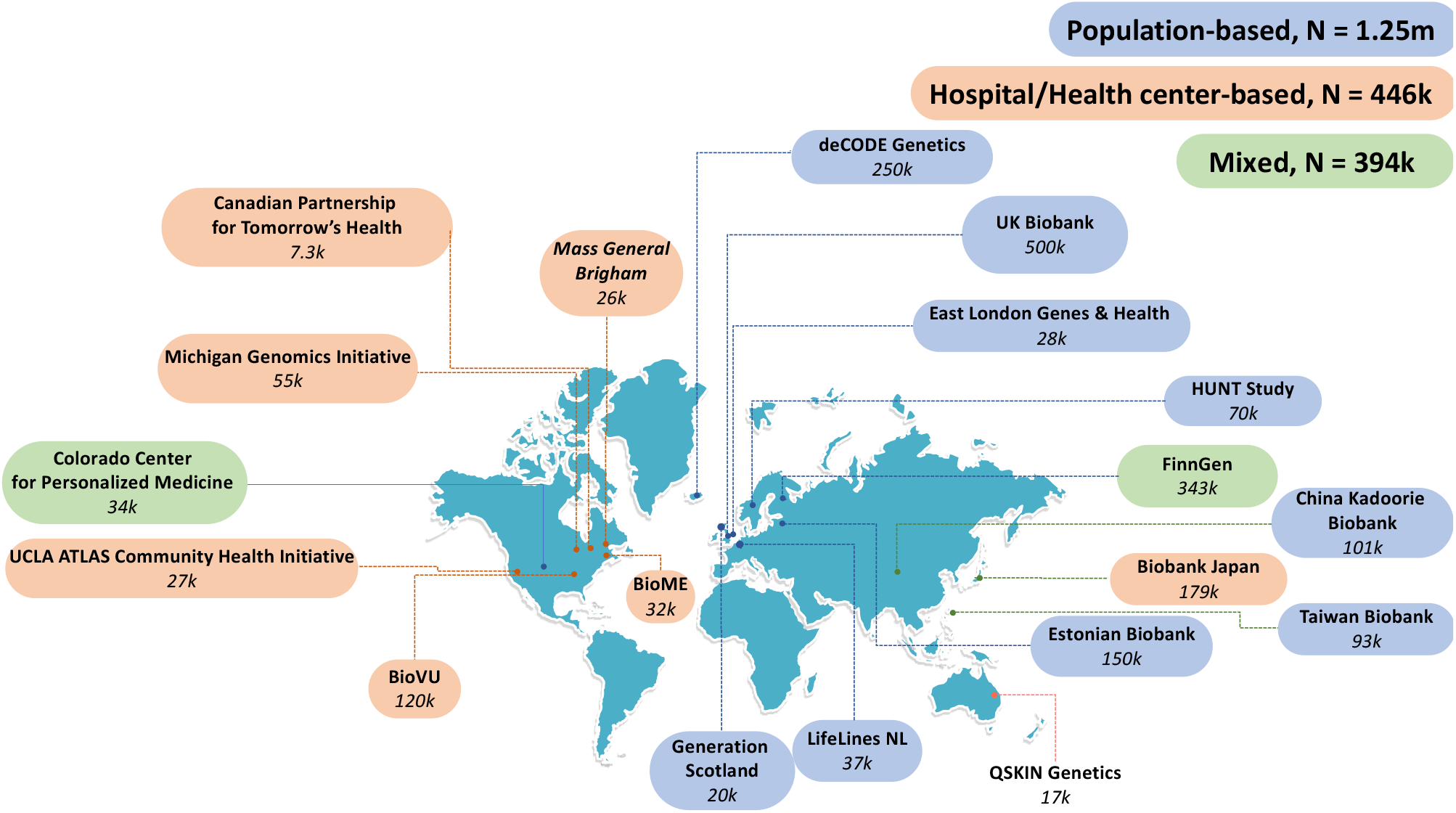
19 biobanks across four continents have joined in GBMI as of November 2021, bringing the total number of samples with matched health data and genotypes to more than 2.1 million. Biobanks are colored based on the sample recruiting strategies.

**At the heart of the GBMI is a community of investigators that have adopted seven principles to guide our collaboration:**

1. **Collaborate in an environment of honesty, fairness and trust;**
2. **Promote early-career researchers;**
3. **Respect other groups’ data;**
4. **Operate transparently with a goal of no surprises;**
5. **Seek permission from each group to use results prior to public release;**
6. **Do not share another group’s results with other parties without permission;**
7. **Do not inhibit any work being done within an individual biobank (or between pairs of biobanks)**.

Here we present the pilot effort of GBMI, in which we meta-analyzed GWAS results for 14 endpoints of common interest (**Supplementary Table 2**). These include diseases across a more than 30x prevalence range: asthma (153,763 cases (sample prevalence: 8.54%)), chronic obstructive pulmonary disease (COPD, 81,568 cases (5.86%)), heart failure (HF, 68,408 cases (5.05%)), and stroke (60,176 cases (4.39%)), gout (37,105 cases (2.50%)), venous thromboembolism (VTE, 27,987 cases (2.63%)), primary open-angle glaucoma (POAG, 26,848 cases (1.80%)), abdominal aortic aneurysm (AAA, 9,453 cases (0.65%)), idiopathic pulmonary fibrosis (IPF, 8,006 cases (0.64%)), thyroid cancer (ThC, 6,699 cases (0.41%)), and cardiomyopathy (HCM, 2,993 cases (0.25%)), a female-specific disease: uterine cancer (UtC, 8,295 cases (1.2%)), and to examine procedure-related phenotypes: acute appendicitis (AcApp, 32,706 cases (2.95%)) and the related appendectomy procedure (14,446 cases (1.86%)), which is an endpoint phenotype that can be extracted from EHR procedure codes but has not been widely studied in previous GWAS. As a proof of concept, using aligned phenotype definitions, analysis methods, sharing standards, and quality control, we demonstrate the advantages of aggregating biobanks together for genetic studies of human diseases.

## Results

### Overview of biobanks in GBMI

GBMI represents 2.1 million research participants with health and genetic data from nineteen biobanks across four continents: one from Australia, three from East Asian countries, eight from European countries, and seven ascertained in North America. Specifically, the biobanks included were: QSKIN Genetics (QSkin)(Olsen et al., 2012) in Australia, Biobank Japan (BBJ)(Nagai et al., 2017), China Kadoorie Biobank (CKB)(Chen et al., 2011), and Taiwan Biobank (TWB)(Feng et al., 2021) in East Asia, deCODE Genetics (deCode)(Gudbjartsson et al., 2015), Estonian Biobank (ESTBB)(Leitsalu et al., 2015), FinnGen, Generation Scotland (GS)(Smith et al., 2013), Genes & Health (GNH)(Finer et al., 2020), LifeLines(Scholtens et al., 2015), Trøndelag Health Study (HUNT)(Krokstad et al., 2013), and UK Biobank (UKBB)(Bycroft et al., 2018) in Europe, and BioME(Abul-Husn et al., 2021), BioVU(Bowton et al., 2015), Canadian Partnership for Tomorrow’s Health (CanPath)(Dummer et al., 2018), Colorado Center for Personalized Medicine Biobank (CCPM)(Aquilante et al., 2020), Mass General Brigham Biobank (MGB)(Karlson et al., 2016), Michigan Genomics Initiative (MGI) (Zawistowski et al., 2021), and UCLA ATLAS Community Health Initiative (UCLA)(Johnson et al., 2021) in North America. **Supplementary Table 1** presents a brief summary of the biobanks in GBMI, including basic information about each biobank (location, institute, cohort size, and sample recruiting approach), participants (ancestry and age), types of electronic health data (self-report data from epidemiological survey questionnaires, billing codes, doctors’ narrative notes, and death registry, etc.) and genotypes (genotyping platforms and imputation reference), as well as data access and references (webpage if available).

Biobanks differ in many aspects, such as locations, sample sizes, genotyping and phenotyping approaches, follow-up time (longitudinal data and samples), and strategies to recruit participants: community/population-based, health center/hospital-based, or mixed (**Figure 1**). As a result, disease prevalence varies across biobanks (**Supplementary Figure 1**) and across sample recruiting strategy groups (**Supplementary Figure 2A**). Biobanks recruiting participants from health centers or hospitals, relative to those recruiting participants from the general population, had a significantly higher prevalence (Wilcoxon test p-value < 0.05) for six out of 13 examined diseases (Appendectomy was excluded from the test due to insufficient data shared from the hospital-based biobanks) (**Supplementary Figure 2B**), including asthma, heart failure (HF), stroke, venous thromboembolism (VTE), gout, and Idiopathic pulmonary fibrosis (IPF).

GBMI incorporates diverse ancestries in genetic studies by including biobank samples of 6 main ancestry groups: approximately 33,000 of African ancestries either from Africa or from admixed-ancestry diaspora (AFR), 18,000 admixed American (AMR), 31,000 Central and South Asian (CSA), 341,000 East Asian (EAS), 1.4 million European (EUR), and 1,600 Middle Eastern (MID) individuals. To compare the ancestries represented between the different biobanks, we projected biobanks’ participants to the same principal components (PC) space (**Figure 2**) using the pre-computed loadings of genetic markers overlapping in all biobanks and the reference containing 1000 Genomes (1000 Genomes Project Consortium et al., 2015) and Human Genome Diversity Project (HGDP) (Cann et al., 2002). PCs projected in the same space enables the cross-comparison of the sample ancestry among all biobanks (**Methods)**. Notably, the population labels used in GBMI were defined by global genetic reference datasets, but GBMI is not globally representative; for example, given that the majority of individuals assigned to AMR and AFR ancestry groups are mostly from biobanks in the US, GBMI participants’ ancestries are not currently representative of broader Central/South American or continental African ancestries, respectively.

**Figure 2.**
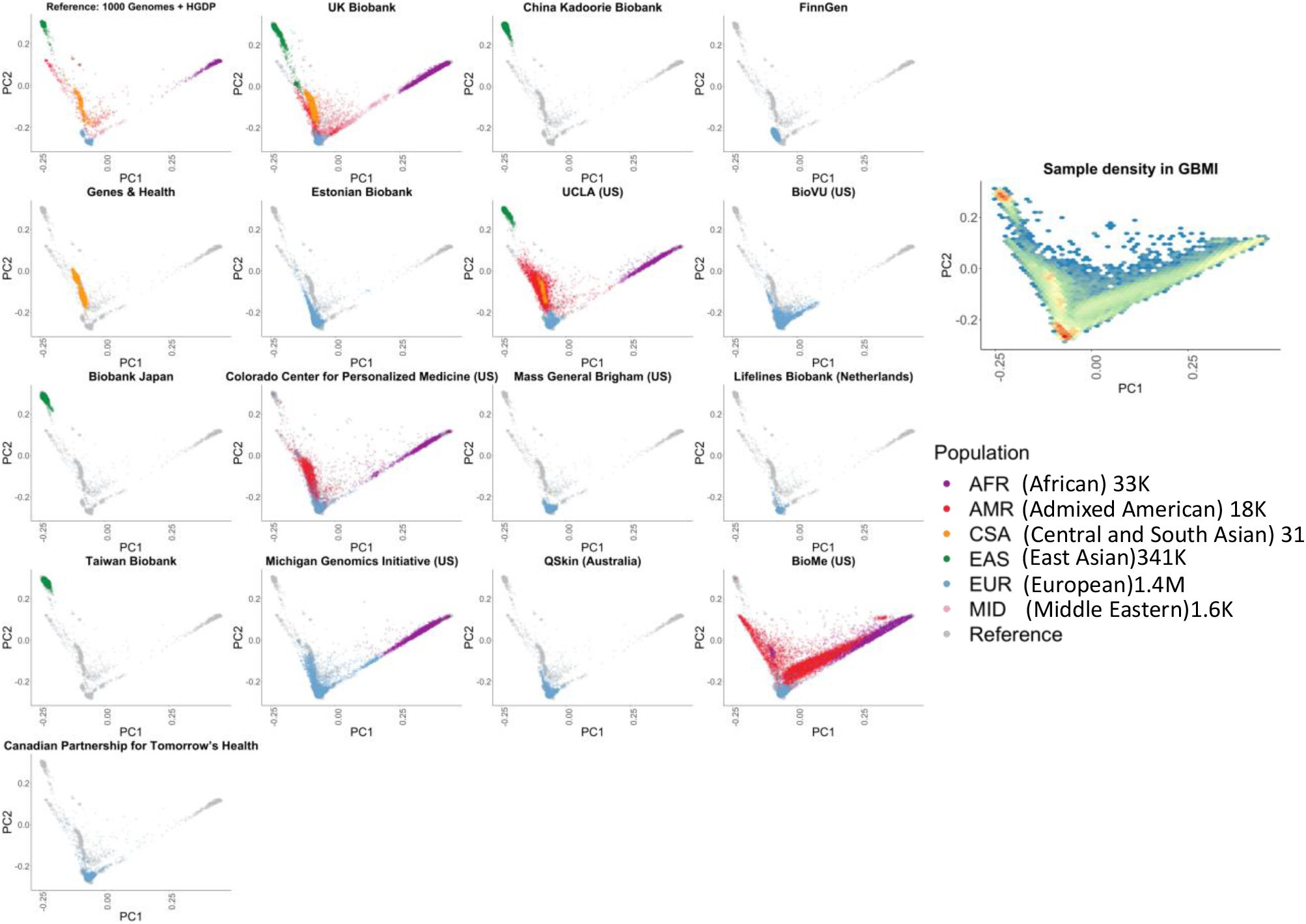
GBMI incorporates biobanks with diverse sample ancestry into genetic studies. Biobanks’ participants were projected to the same principal components (PC) space using the pre-computed loadings of genetic markers.

### All-Biobank Meta-analysis

The overview of the meta-analysis is presented in **Supplementary Figure 3**. We harmonized the phenotype definitions primarily by mapping the International Classification of Diseases (ICD) codes to phecodes (Denny et al., 2013) for diseases and Classification of Interventions and Procedures codes (OPCS) for procedures, which were used by biobanks to curate phenotypes based on the health data available (**Supplementary Table 3** and **4**). After standard sample-level and variant-level quality control (**Supplementary Table 1**), GWASs stratified by ancestry and sex were conducted in each biobank. The central analysis team performed post-GWAS variant-level quality control for each biobank by flagging markers with different allele frequencies compared to gnomAD (Karczewski et al., 2020) and excluding markers with imputation quality score < 0.3 (**Methods**). Across all biobanks, 70.8 million genetic variants were tested for associations, of which 2.9 million variants are protein coding (**Supplementary Table 5**).

We first highlight the power of GBMI to uncover novel genetic associations through increased sample size. Inverse variance-based meta-analysis of all biobanks for the 14 endpoints successfully replicated 320 previously reported loci and identified 188 apparently novel loci, spanning the frequency spectrum of less frequent to common variants (**Supplementary Table 6, Figure 3**). At 87 loci, a protein coding variant was the most significant variant (n=26) (**Table 1**) or in linkage disequilibrium with the most significant variant with r^2^ > 0.8 (n=66 additional) at the locus. 18 of these 87 loci are novel (**Supplementary Table 7**). Furthermore, 13 out of 14 endpoints have SNP-based heritability significantly different from 0 on the liability scale, (under the assumption that the population prevalence matches the prevalence of all biobanks aggregated together), ranging from 1.79% (acute appendicitis) to 10.73% (Gout) (**Supplementary Table 8**).

**Table 1.** Lead variants that are protein coding within 26 disease-associated loci identified in the multi-biobank multi-ancestry meta-analyses in GBMI

| Endpoint | CHR/POS (hg38) | REF/ALT | Freq<sup>a</sup> | Odds Ratio (95% CI)<sup>b</sup> | p-value | Heterogeneity p-value | Gene | Function | cases | controls | Number of biobanks |
| --- | --- | --- | --- | --- | --- | --- | --- | --- | --- | --- | --- |
| **Novel** |  |  |  |  |  |  |  |  |  |  |  |
| AAA | 10:73913343 | T/C | 0.737 | 0.88 (0.85-0.91) | 5.93E-12 | 0.76 | *PLAU* | missense | 9,453 | 1,446,422 | 11 |
| COPD | 1:149934520 | T/C | 0.350 | 1.04 (1.03-1.05) | 7.91E-10 | 0.54 | *MTMR11* | missense | 79,844 | 1,289,683 | 15 |
| Stroke | 6:49492265 | A/G | 0.446 | 0.96 (0.94-0.97) | 1.8E-11 | 0.99 | *CENPQ* | missense | 60,176 | 1,310,725 | 16 |
| Asthma | 10:94279840 | G/C | 0.448 | 1.03 (1.02-1.03) | 2.52E-09 | 0.98 | *PLCE1* | missense | 153,763 | 1,647,022 | 18 |
| Asthma | 14:100883117 | G/T | 0.025 | 1.09 (1.05-1.12) | 2.61E-08 | 0.73 | *RTL1* | missense | 133,369 | 1,370,606 | 16 |
| Asthma | 19:56222056 | C/A | 0.253 | 1.03 (1.02-1.04) | 2.35E-08 | 0.60 | *ZSCAN5A* | missense | 149,293 | 1,626,581 | 17 |
| **Known** |  |  |  |  |  |  |  |  |  |  |  |
| COPD | 14:94378610 | C/T | 0.020 | 1.22 (1.16-1.29) | 5.2E-15 | 9.27E-03 | *SERPINA1* | missense | 54,105 | 883,399 | 11 |
| COPD | 19:44908684 | T/C | 0.140 | 0.95 (0.94-0.97) | 1.04E-08 | 0.36 | *APOE* | missense | 81,568 | 1,310,798 | 16 |
| Gout | 2:27508073 | T/C | 0.588 | 0.87 (0.86-0.88) | 9.27E-64 | 0.11 | *GCKR* | missense | 37,105 | 1,448,128 | 15 |
| Gout | 11:64593747 | G/A | 0.016 | 0.36 (0.31-0.42) | 1.19E-41 | 0.10 | *SLC22A12* | stop gain | 6,634 | 248,305 | 2 |
| Gout | 12:57449928 | G/A | 0.194 | 0.91 (0.89-0.93) | 1.51E-17 | 0.45 | *INHBC* | missense | 37,105 | 1,448,128 | 15 |
| IPF | 5:1279370 | T/C | 0.001 | 862 (205-3618) | 2.66E-20 | 0.09 | *TERT* | missense | 1,278 | 330,954 | 2 |
| IPF | 5:169588475 | G/A | 0.014 | 2.19 (1.81-2.66) | 1.61E-15 | 0.02 | *SPDL1* | missense | 4,812 | 882,416 | 7 |
| POAG | 1:11193760 | C/T | 0.026 | 0.67 (0.6-0.74) | 4.39E-14 | 0.90 | *ANGPTL7* | missense | 12,810 | 421,360 | 5 |
| POAG | 1:171636338 | G/A | 0.002 | 6.33 (4.71-8.51) | 1.67E-34 | 4.33E-06 | *MYOC* | stop gain | 15,916 | 1,092,446 | 11 |
| POAG | 14:60509819 | C/A | 0.547 | 0.89 (0.87-0.91) | 7.08E-30 | 0.31 | *SIX6* | missense | 26,848 | 1,460,599 | 15 |
| Stroke | 12:111803962 | G/A | 0.238 | 0.9 (0.88-0.92) | 5.16E-18 | 0.37 | *ALDH2* | missense | 23,804 | 269,656 | 4 |
| VTE | 1:169549811 | C/T | 0.020 | 3.04 (2.85-3.24) | 1.5E-245 | 5.52E-13 | *F5* | missense | 26,749 | 1,011,509 | 9 |
| VTE | 12:6034818 | T/C | 0.889 | 1.1 (1.07-1.13) | 1.59E-10 | 0.21 | *VWF* | missense | 27,987 | 1,035,290 | 9 |
| VTE | 12:103742510 | C/T | 0.011 | 1.65 (1.45-1.88) | 6.19E-14 | 0.25 | *STAB2* | missense | 10,353 | 341,418 | 2 |
| Asthma | 1:12115601 | G/A | 0.012 | 0.85 (0.8-0.89) | 1.7E-11 | 0.46 | *TNFRSF8* | missense | 118,767 | 1,202,660 | 12 |
| Asthma | 1:31699894 | G/T | 0.573 | 1.03 (1.02-1.04) | 1.61E-10 | 0.49 | *COL16A1* | missense | 148,045 | 1,579,632 | 17 |
| Asthma | 4:38797027 | C/A | 0.387 | 0.95 (0.94-0.96) | 4.21E-21 | 0.54 | *TLR1* | missense | 138,764 | 1,458,022 | 15 |
| Asthma | 4:102267552 | C/T | 0.044 | 1.08 (1.06-1.1) | 2.53E-12 | 0.81 | *SLC39A8* | missense | 129,434 | 1,256,670 | 14 |
| Asthma | 5:14610200 | C/G | 0.084 | 1.07 (1.05-1.09) | 7.68E-15 | 0.16 | *OTULINL* | missense | 125,483 | 1,241,068 | 13 |
| Asthma | 9:128721272 | T/A | 0.068 | 0.95 (0.93-0.96) | 5.61E-10 | 0.21 | *ZDHHC12* | missense | 152,469 | 1,638,824 | 18 |

<sup>a</sup>Frequencies are reported with respect to the alternate allele (ALT) in the combined meta-analysis data sets.
<sup>b</sup>Odds ratios are reported with respect to the alternate allele (ALT) in the meta-analyses.

**Figure 3:**
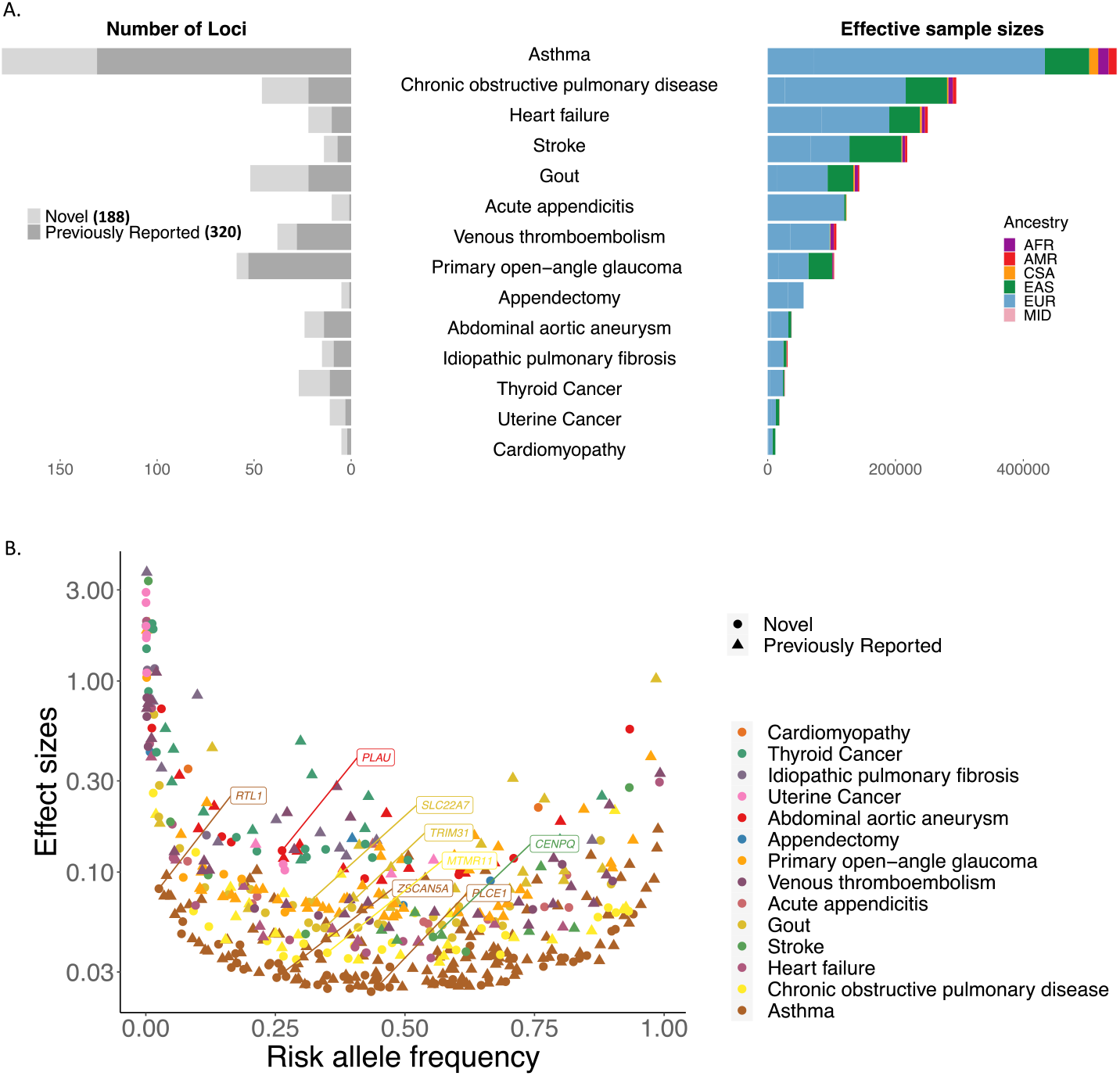
All-biobank meta-analysis for the 14 endpoints have successfully replicated 320 previously reported loci and identified 188 novel loci. A. number of loci were plotted for each endpoint (left panel) against the effective sample sizes 1/(4/cases + 4/controls) colored by the sample ancestry (right panel). B. Top hits spread over the entire allele frequency spectrum. Phenotypes are in ascending order by the effective sample sizes. 3 markers with beta > 5 are not shown. Gene names are labeled for the novel loci with protein-coding index variants.

Secondly, we show that identified associations were largely shared across biobanks. The lead variants at ∼96% of the 508 genome-wide loci did not show evidence for heterogeneity in effect sizes across different data sets (per biobank and ancestry) (**Supplementary Table 6**) with p-value for Cochran’s Q test ≥ 1/508, despite biobanks differing in many aspects, as discussed above. This suggests that harmonizing phenotyping and then integrating GWASs from different biobanks together with the analysis pipeline within GBMI enables reliable discoveries for genetic-disease associations. Out of the lead variants for 27 loci showing evidence for heterogeneity in effect sizes, 11 have heterogeneous effect sizes across ancestry groups (**Supplementary Table 6**). We additionally used the meta-regression approach implemented in MR-MEGA (Mägi et al., 2017) to conduct the all-biobank meta-analysis across all ancestries. In contrast with a fixed-effects, inverse variance-based meta-analysis, MR-MEGA accounts for the effect size heterogeneity across data sets. This led to the identification of 17 additional loci across 10 endpoints, including 12 that were novel (**Supplementary Figure 4, Supplementary Table 9**).

#### Power improved by incorporating samples with non-European ancestries

An additional 21.8 million genetic variants were analyzed in the all-biobank meta-analysis which were not present in the European HRC-imputed variant sets. The majority of these variants were rare, with 18 million having a MAF ≤ 1%, and the other 3.8 million were common in at least one ancestry group (**Supplementary Figure 5**). Incorporating samples with diverse ancestries to the biobank meta-analysis enabled the comparison of effect sizes of genomic loci across ancestry at associated loci. Nine out of the 499 loci that were tested in more than one ancestry showed evidence for heterogeneity in effect sizes across ancestry (p-value for Cochran’s Q test across ancestry < 1/499) (**Supplementary Table 10**). 343 loci were identified in the European-only meta-analysis and the inclusion of the non-European samples yielded an additional 165 loci (**Supplementary Figure 6A, Supplementary Table 11)**, bringing the total number of loci to 508 in the all-biobank meta-analysis. While increase in sample size will drive some of this increase alone, the increased diversity allows to identify loci whose index variants are much more frequent in other ancestries than in European ancestries. In particular, in contrast to only 4 out of the 343 loci (1.16%), 21 out of the 165 additional loci (12.7%) have index variants that are at least 10 times more frequent in other ancestries than in European ancestries with the frequency lower than 5% in European ancestries (**Supplementary Table 11)**. Forest plots of several example loci are shown in **Supplementary Figure 6B**, highlighting loci whose index variants that are more frequent in East Asians than Europeans and other ancestries (*MIR2054/INTU* for POAG, *PNPT1/EFEMP1* for COPD, and *NAA38* for Asthma) as well as loci driven by variants that are more frequent in African ancestry than other ancestries, including *VPS13D/DHRS3* for VTE, *BCL2L12* for HF, and *MEIS2/TMCO5A* for stroke.

#### Sex-stratified meta-analyses

We performed sex-stratified meta-analysis to compare GWAS effect sizes between sexes. 479 loci were tested in more than one biobank for both male-only and female-only meta-analyses, of which eight loci showed evidence of heterogeneous effect sizes between males and females (p-value for Cochran’s Q test < 1/479) in the all-biobank meta-analysis (**Supplementary Table 12**). This included one novel locus for the studied diseases: a region containing *CTDP1/KCNG2* for asthma (**Supplementary Figure 7A**) and seven previously identified loci (**Supplementary Figure 7B**).

Environmental factors, such as smoking status and alcohol usage, that are different in males and females may play a role in GWAS effect size differences among sexes. For example, the most strongly associated variant in the *CTDP1/KCNG2* locus (rs11665567) is an intergenic variant with a female-specific association for asthma (in females allele frequency(AF) = 18.8%, effect size (se) = 0.05 (0.008), p-value = 5.62×10^−10^; in males AF=18.7%, effect size (se) = 0.003 (0.01), p-value = 0.75 (p-value for difference=2.4×10^−4^).This locus was previously shown to be associated with smoking status (Liu M et. al. 2019). In addition, we have replicated two previously reported loci that are located at the aldehyde dehydrogenase family genes for gout exhibiting stronger associations in males than in females (Mizuno et al., 2015; Sulem et al., 2011). One is the East Asian-specific intronic variant rs4646776 (r^2^ = 0.99 with the missense variant rs671(Matoba et al., 2020)) located at the gene *ALDH2* with a stronger effect in males than in females (in females AF = 20.4%, effect size (se) = -0.10 (0.056), p-value = 0.07, in males AF=24.2%, effect size (se) = -0.29(0.023), p-value = 2.5×10^−36^). This has been attributed to the higher frequency of habitual alcohol drinking in males than females among individuals of East Asian ancestries (Mizuno et al., 2015). The other one is the low-frequency European-specific intronic variant located in *ALDH16A1* that is associated with serum urate levels (Sulem et al., 2011) more strongly in men(rs752383928 intronic, in females AF = 0.74%, effect size (se) = 1.63 (0.29), p-value = 2.43×10^−8^, in males AF=0.73%, effect size (se) = 2.70 (0.18), p-value = 1.33×10^−50^). To clarify whether the sex-specific associations identified are due to pleiotropic effects of the genetic variants, environmental factors, or possible gene-environment interactions requires further study.

In addition, we uncovered significant sex differences for five previously reported loci: *RANBP6/IL33* for asthma(Demenais et al., 2017), *AFAP1* for COPD(Wyss et al., 2018), *PKD2* for gout (Matsuo et al., 2016), *MUC5AC/MUC5B* for IPF(Seibold et al., 2011), and *ARHGEF12* for POAG (Springelkamp et al., 2015).

Furthermore, there were 31 loci only identified in the sex-stratified meta-analyses but not in the sex-combined meta-analyses (p-value > 5×10^−8^), of which 11 loci were detected in female-only meta-analyses and 20 loci only in male-only meta-analyses. 26 out of the 31 loci are potentially novel for the studied phenotypes (**Supplementary Table 13**). The female-only meta-analysis for stroke identified the previously reported locus *CETP* (Buraczynska et al., 2018) that did not reach the genome-wide significance threshold in the sex-combined meta-analysis. The top hit is an intronic rs7499892 variant with stronger association in females than males (in females: effect size (se) = 0.078 (0.014), p-value =1.08×10^−8^, in males effect size (se) = 0.007(0.012), p-value = 0.56). Previous studies have shown that the transgenic expression of *CETP* increases plasma triglyceride levels in females and males through distinct mechanisms (Palmisano et al., 2016, 2021).

### Integration of association results across biobanks

Leveraging the heterogeneity among biobanks, we used the genetic association results to evaluate the integration of biobanks in the meta-analyses for genetic discovery. First, we compared the effect sizes of the index variants with p-value < 1×10^−10^ by the all-biobank meta-analysis in each biobank and leave-one-biobank-out meta-analysis (LOBO). For the biobank and LOBO pairs, we fit a Deming regression model (Deming, 1943), which accounts for the standard errors of effect sizes in both association datasets, with the intercept set to zero. In **Supplementary Figure 8**, the slope estimates for the biobank and LOBO pairs were plotted against the effective sample sizes with biobanks annotated by phenotype source (health records (e.g. ICD codes and physician’s diagnosis) only or self-reported data included), sampling strategy, and sample ancestry. Most of the slope estimates were not significantly different from one across biobanks and phenotypes suggesting the genetic association results are robust despite differences between biobanks. However, we observed exceptions to this among biobanks with relatively smaller sample sizes and/or non-European or multiple ancestries. For example, the multi-ancestry biobanks BioMe, BioVU, and UCLA as well as GNH, which included SAS samples, have different effects compared to others for multiple phenotypes, including gout, HF, VTE, and POAG. Note that POAG tends to have more phenotypic heterogeneity due to glaucoma types not being well defined by self-reported data, leading to the inclusion of other types of glaucoma, such as the angle-closure glaucoma. As expected, the three biobanks using self-reported data for phenotype curation, Lifelines, TWB, and BBJ, also showed effect size differences for POAG compared to other biobanks (**Supplementary Figure 8**).

Next, we estimated the genetic correlation between individual biobanks and LOBO for the three endpoints with the highest heritability estimates: asthma, gout, and COPD. Genetic correlation estimates between biobanks and the LOBO were close to 1, although genetic correlation was only possible to estimate for biobanks with non-zero heritability estimates (p-value < 0.05) (**Methods**) (**Supplementary Figure 9**). We then compared the all-biobank meta-analyses with previously published GWAS studies. Among previously reported loci, we show consistent effect directions between GBMI and the previous largest studies. For example, all 18 loci for asthma that were previously identified by the Trans-National Asthma Genetic Consortium (TAGC) (Demenais et al., 2017) have consistent effect directions in GBMI. Similarly, 24 out of 25 previously identified loci by MVP for AAA (Klarin et al., 2020) and 40 out of 40 previously identified loci for gout (Tin et al., 2019) show effect size consistency between GBMI and the previous GWASs (**Supplementary Table 14**). Note that by cross-comparing the cohort lists in previous studies and GBMI, no sample overlap was noted for Asthma and AAA, while 3 biobanks in GBMI (BioVU, GS, and UKBB) were also included in the previous meta-analysis for gout (Tin et al., 2019), accounting for about 20% of the GBMI samples.

### Biological implications of genetic associations

#### Pleiotropic effects of associated loci

We investigated the genetic relationship between the endpoints and other complex traits by examining the associations of the top variants identified by the all-biobank meta-analyses with 1,238 human diseases in UKBB (**Methods**). Of the 430 loci whose index variants were available in the UKBB GWAS data, 78 variants identified from 13 GBMI endpoints (except for HCM) exhibited significant (p-value < 5 × 10^−8^) pleiotropic associations with at least one other phenotype (**Supplementary Table 15**). Risk increasing alleles of the top variants at two asthma-associated loci, the known asthma locus *BACH2* and the novel locus *FGFR1OP*, are both associated with a reduced risk of hypothyroidism. The risk increasing allele of the top variant at the novel locus *GOT1/LINC01475* for acute appendicitis (AcApp) is associated with the decreased risk of ulcerative colitis. A previous study also observed a low risk of ulcerative colitis among people who had undergone an appendectomy for appendicitis and mesenteric lymphadenitis(Andersson et al., 2001), but the reason for this remains unclear.

#### Prioritization of cell types, tissues, and genes

To further understand the biology underlying the genetic associations, we first prioritized the tissues and cell types in which genes at the associated loci are likely to be highly expressed using the Data-driven Expression-Prioritized Integration for Complex Traits (DEPICT)(Pers et al., 2015) (**Supplementary Table 16)**. For example, at FDR < 0.05, the adrenal cortex, which releases the mineralocorticoid aldosterone, was prioritized for AAA consistent with previous functional studies which have shown that the mineralocorticoid aldosterone can induce aortic aneurysm and dissection in the presence of high salt(Liu et al., 2013). Prioritized tissue types for asthma included lymphoid tissue and immune systems (blood cells, antigen presenting cells, and myeloid cells) as well as nasal and respiratory mucosa. Besides muscle cells and connective tissue cells, heart and blood vessels were identified for POAG(Lo Faro et al., 2021).

Next, several methods were used to prioritize the potentially functional genes, including DEPICT (**Supplementary Table 17)**, the gene-level Polygenic Priority Score (PoPS) (Weeks et al., 2020) (**Supplementary Table 18)**, transcriptome-wide association studies (TWAS) (Bhattacharya et al., 2021a) (**Supplementary Table 19)**, and proteome-wide Mendelian randomization (PWMR) (Zhao et al., 2021) (**Methods**). Using asthma, POAG, and VTE as examples, the gene lists generated by these different methods showed quite little overlap (**Supplementary Figure 10**). For asthma, 618 genes were prioritized by at least one of the four approaches (FDR < 0.05 by DEPICT, top 1% scores in PoPS, P < 2.5×10^−6^ by TWAS, P < 0.001 by PWMR) (**Supplementary Figure 10A**). However no genes were prioritized by all four methods and 5 were prioritized by any three out of the four methods (*FCER1G, IL18R1, IL4R*, and *SMAD3* by DEPICT, TWAS, and PoPS and *IL2RB* by DEPICT, PoPS, and PWMR) (**Supplementary Table 20**). All these genes are located at the well-known asthma-associated loci. *FCER1G* encodes the Fc Fragment of IgE Receptor Ig, and the *IL18R1, IL4R*, and *IL2RB* encode Interleukin receptors, which are all involved in the immune system. Dupilumab, an anti-interleukin 4 receptor alpha monoclonal antibody, blocks IL-4 and IL-13 and decreases IgE over time, is an FDA approved add-on therapy for asthma (Castro et al., 2018; Rabe et al., 2018). *SMAD3* encodes a transcription factor whose methylation has been shown to be associated with neonatal production of IL-1β and childhood asthma risk (DeVries et al., 2017). Similarly, for POAG, 204 genes were prioritized, but no genes were prioritized by all four or any three methods (**Supplementary Figure 10B**) For VTE, 244 genes were prioritized, one well-known VTE-associated gene, *F2*, that encodes the coagulation factor II, was prioritized by all four methods, and 5 genes were prioritized by any three methods (*F5, PLCG2, PLEK, PROC*, and *PROS1* by DEPICT, PWMR, and PoPS) (**Supplementary Figure 10C**). In line with what has previously been discussed (Weeks et al., 2020), these results showed that the existing gene prioritization methods successfully prioritized relevant genes for diseases but have poor agreement. Note that besides adapting different statistical models and pipelines, these approaches prioritize genes based on different expression data types; DEPICT, PoPS, and TWAS are based on expression quantitative trait loci (eQTLs), while PWMR uses the protein quantitative trait loci (pQTLs) (**Methods**). Our results highlight the challenges in interpreting genome-wide significant loci and the clear need for robust in silico approaches and pipelines to nominate genes for experimental follow-up.

### Biobank meta-analysis for genetic association studies

#### Improving power of genetic discovery for common diseases

Aggregating 18 biobanks in GBMI brings a substantial increase in samples sizes for genetic association studies for asthma (153,763 cases and 1,647,022 controls) compared to the previous largest meta-analysis by TAGC (Demenais et al., 2017) with 66 individual asthma cohorts (23,948 cases, 118,538 controls) (**Supplementary Table 2)**. The larger sample size leads to an increase in power for genetic discovery; 180 genome-wide significant loci for asthma were identified by GBMI, of which 49 are novel (**Supplementary Table 6**). Furthermore, all 18 loci that were first reported by TAGC have more significant association p-values in GBMI (**Supplementary Figure 11**). In addition, meta-analyzing GBMI biobanks and the existing disease consortia would further increase the discovery power to uncover genetic risks for human diseases. For example, we meta-analyzed 14 biobanks in GBMI with two previous meta-analysis studies for POAG (excluding three overlapped biobank data sets), which doubled the case numbers compared to the previous largest meta-analysis and successfully identified 103 significant loci, of which 19 are novel (Lo Faro et al., 2021).

#### Providing opportunities for genetic studies on less prevalent diseases

EHR-linked biobanks provide opportunities to assess less prevalent diseases that were understudied by previous GWAS studies. For example, although gout has caused an increased health burden in recent years, the largest meta-analysis for gout so far was conducted on 13,179 cases and 750,634 controls across 20 studies. By meta-analyzing 15 biobanks in GBMI comprising 37,105 cases and 1,448,128 controls with 5 ancestral populations (**Supplementary Table 2**), we have identified 52 significant loci, of which 30 are novel (**Supplementary Table 6**). As expected, a vast majority of these loci (n=40, either same SNP or multiple different SNPs) were associated with serum urate levels (Gill et al., 2021; Huffman et al., 2015; Sinnott-Armstrong et al., 2021; Tin et al., 2019) (**Supplementary Table 21)**, including key urate transporter genes *SLC2A9* and *SLC22A12* (Xu et al., 2017). To further identify the potential biological explanation of the loci with gout risk in our study, we explored their associations with other relevant phenotypes. We find that 30 of these loci are associated with other relevant traits and diseases. For example, *RAB24, STC1*, and *MAF* are associated with BMI, *MPPED2, BCAS3* with kidney function, *HNF4A, PNPLA3, MC4R* with diabetes, *GCKR* with glycolysis and *ARID1A, MLXIPL, A1CF, INHBC* among others are associated with blood pressure and lipids (HDL-C, LDL-C, TG, and Apolipoprotein B). Previous studies have already speculated the possible mechanisms for the involvement of these traits or processes in gout etiology. For example, coating of urate crystals with Apolipoprotein B can down-regulate the innate immune system by suppressing neutrophil activation(Terkeltaub et al., 1984) and neutrophil activation is needed for the endocytosis and lysis of urate crystals and thus the resolution of gout attack (So and Martinon, 2017). Similar biological links have also been proposed for other above-mentioned traits and uric acid metabolism and thus can explain the observed association of related genes with gout risk in our study.

Biobanks also enable genetic studies of different types of disease phenotypes. We conducted biobank meta-analysis for acute appendicitis and the relevant procedure endpoint appendectomy and observed high genetic correlation between the two endpoints (r^2^ = 0.99). Out of the 10 loci identified for acute appendicitis by meta-analyzing 10 biobanks, 3 were also significant for appendectomy (**Supplementary Table 6**) even though the sample size was 3 times lower, suggesting that the procedure phenotypes may add meaningful information in biobank-based genetic studies and incorporating these phenotypes to traditional disease diagnosis phenotypes could improve discovery power.

#### Improving polygenic risk scores based on multi-biobank multi-ancestry meta-analyses

Based on the leave-one-biobank-out meta-analyses in GBMI, biobanks can estimate the predictive value of PRS for the endpoints. Using asthma as an example, we have demonstrated the improved PRS prediction based on the GBMI summary statistics compared to the previous meta-analysis by TAGC (Demenais et al., 2017) in 6 biobanks across 6 ancestral populations (**Supplementary Figure 12**). Wang et al has evaluated the performance of PRS using the multi-biobank multi-ancestry meta-analyses of GBMI for additional endpoints and demonstrated improved PRS prediction in individual biobanks based on GBMI results compared to previous GWASs. The Bayesian method PRS-CS(Ge et al., 2019) overall outperformed the classic pruning and thresholding method, P+T, especially for endpoints with higher SNP-based heritability, while the PRS prediction accuracy varies across biobanks and ancestries. The EUR-based LD reference panel provides comparable or better prediction accuracy relative to using other cosmopolitan LD panels for PRS construction methods based on GBMI association results as the EUR ancestry constitutes the largest proportion (about 69%) of GWAS participants (Wang et al., 2021).

## Discussion

Genetic discovery benefits from the increasing numbers of EHR-linked biobanks, despite biobanks differing in many aspects. As of September 2021, 19 biobanks across four continents comprising six major ancestral groups have joined GBMI for the globally collaborative efforts to uncover genetic risk factors of human diseases. As the pilot effort in GBMI, with carefully harmonized phenotype definitions and analysis pipelines, we meta-analyzed GWASs in up to 18 biobanks for 14 endpoints, including both common diseases (asthma, COPD, VTE, etc.) and less prevalent diseases (gout, IPF, AAA, thyroid cancer, etc.). Over 500 genome-wide significant loci were detected of which 188 are novel. Sex-stratified meta-analysis allows for comparing effects between sexes and identified 8 loci with different effect sizes in men and women. Besides demonstrating the integration of genetic association results from different biobanks, our results have illustrated the gains by meta-analyzing biobanks together. The increase in the sample sizes and sample diversity leads to higher discovery power. Incorporating non-European samples in the meta-analysis allows for the genetic association tests for 21.8 million additional markers, of which more than 85% are low-frequency variants (AF < 1%), which may facilitate functional follow-up studies to disentangle the causal variants at identified loci.

Gout has a prevalence of 1-4% worldwide and affects more than 8 million people in the United States (Zhu et al., 2011), but has been less well studied by previous GWASs. Meta-analyzing biobanks increases the case number of gout by three times compared to the previous GWAS, which uncovered 30 novel loci. In addition, biobank meta-analyses were done for different types of endpoints that can be accessed through EHRs in biobanks but not studied by previous GWASs, such as the procedure endpoint appendectomy and the relevant disease acute appendicitis. The high genetic correlation between the two endpoints suggests potential usage of the non-disease endpoints derived from EHRs for genetic discovery. As expected, based on the leave-one-biobank meta-analysis results, more accurate predictive polygenic risk scores, especially for biobank samples with European and East Asian ancestries, were constructed due to the increased genetic discovery power compared to previous studies. This gain can be further extended to non-European samples as the sample diversity continues to increase in GBMI. The collaborative efforts of biobanks in GBMI creates invaluable resources and opportunities to advance the understanding of the etiology of human diseases, leading to better treatment and prevention, and helps move toward the equitability of genetic studies in diverse ancestries.

We formed multiple working groups to 1. deepen the genetic investigation of the biological implications of results for several endpoints, including asthma (Tsuo et al., 2021), COPD (Tsuo et al., 2021), VTE (Wolford et al., 2021), POAG (Lo Faro et al., 2021), stroke (Surakka et al., 2021), and heart failure (Wu et al., 2021), 2. systematically characterize genome-wide significant loci via fine-mapping(Kanai et al., 2021), transcriptome-wide association (Bhattacharya et al., 2021a), protein QTL Mendelian randomization analysis (Zhao et al., 2021), prioritizing drug targets (Namba et al., 2021) (Namba et al., 2021), and improving the disease risk prediction by polygenic risk scores based on the multi-biobank multi-ancestry meta-analysis results (Wang et al., 2021).

Together, the pilot work conducted in GBMI has shown that despite the heterogeneities across biobanks in many aspects, such as locations, sample sizes, genotyping and phenotyping approaches, sample ancestries, and strategies to recruit participants, with standardized phenotype definitions and the analysis pipeline, biobanks can be meta-analyzed together to provide reliable genetic discoveries. Biobank meta-analysis can have substantial benefits to advance the genetic discoveries for human diseases with the larger sample sizes and the increased ancestry diversity. We have evaluated the challenges in multiple down-stream in silico studies that are used for prioritizing the functional genes and variants, provided the best practices and pipelines based on our lessons from GBMI, and highlighted the need for new method development to address the upcoming issues in the current analysis based on the biobank meta-analysis results.

### Code and Data availability

The all-biobank meta-analysis results and plots for the 14 endpoints (including both ancestry-specific and cross-ancestry meta-analyses and sex stratified meta-analyses) are available for downloading at https://www.globalbiobankmeta.org/resources and browsed at the browser http://results.globalbiobankmeta.org. Custom scripts used for quality control, meta-analysis and summary of results are available at https://github.com/globalbiobankmeta. The optimized trans-ancestry and single-ancestry polygenic score weights will be deposited within the PGS Catalog (https://www.pgscatalog.org/).

## Methods

### Phenotype definition

A phenotype definition guideline was created and shared with all biobanks (**Supplementary Table 3**). The disease endpoints were defined following the phecode maps (Denny et al., 2013), which maps the ICD-9 or ICD-10 codes into hierarchical phecodes, each representing a specific disease group. Study participants were labeled a phecode if they had one or more of the phecode -specific ICD-9 or ICD-10 codes. Cases were all study participants with the phecode of interest and controls were all study participants without the phecode of interest or any related phecodes. For sex-specfic disease endpoint, which is uterine cancer (UtC) in the endpoint list, only females were included in the study samples. The procedure endpoint, appendectomy, was defined based on the OPCS. Any biobank participant with codes H01 (Emergency excision of appendix), H01.1(emergency excision of abnormal appendix and drainage), or H01.2 (emergency excision of a normal appendix) were cases, all other participants without these codes were controls. Biobanks which do not collect the ICD codes or OPCS codes define the phenotypes using the available EHRs according to the phenotype definitions in the guideline.

### GWAS

Each biobank conducted genotyping, imputation, and quality controls independently, followed by running GWASs following the analysis plan shared in GBMI (information available at https://www.globalbiobankmeta.org/) with phenotypes curated according to the harmonized phenotype definitions (see the Phenotype definition section in Methods). We recommended to run GWAS analysis using Scalable and Accurate Implementation of GEneralized mixed model (SAIGE)(Zhou et al., 2018) or REGENIE (Mbatchou et al., 2021), which are scalable for biobank-scale data and account for sample relatedness and case-control imbalances. The suggested covariates were age, age2, sex, age*sex, 20 first principal components, and any biobank specific covariates, such as genotyping batches and recruiting centers.

### Post-GWAS Quality control

Variant-level quality control was conducted for each data set containing GWAS summary statistics shared by biobanks (**Supplementary Table 22, Supplementary Figure 13 and 14**). Genetic variants with MAC < 20 and variants that are poorly imputed with an imputation score < 0.3 were firstly excluded. Genome coordinates of all genetic variants were lifted to GRCh38. For palindromic SNPs (with A/T or G/C alleles), we compared their allele frequencies of the aligned reference allele in the GWAS data set (AF-GWAS) to gnomAD (Karczewski et al., 2020) (AF-gnomAD) by ancestry. If a palindromic SNP meets any one of the following standards, we flip its alleles in the GWAS data set and indicate that this variant has the potential strand flip with a flag: 1. The fold difference is greater than two, 2. The allele frequency of the alternative allele in the GWAS data set is closer to AF-gnomAD than the reference allele, 3. AF-GWAS < 0.4 and AF-gnomAD > 0.6, 4. AF-GWAS > 0.6 and AF-gnomAD < 0.4. We then identified genetic variants with different allele frequencies compared to gnomAD. For each genetic variant, the Mahalanobis distance between AF-GWAS and AF-gnomAD was estimated and the variant was flagged to have different AF-GWAS and AF-gnomAD if the Mahalanobis distance is greater than 3 standard deviations away from the mean. We observed that across 18 biobanks that shared GWAS summary statistics to the meta-analysis for asthma, very small proportions (0.003% to 0.65%) of variants were flagged as either palindromic SNPs with flipped strands or variants having very different allele frequencies compared to gnomAD.

### Meta-analysis

Fixed-effect meta-analyses based on inverse-variance weighting were performed for all endpoints with 1. all biobanks across all ancestries, 2. leave-one-biobank out across all ancestries 3. all biobanks by each ancestry, and 4. all biobanks by sex. Trans-ancestry meta-analysis was performed using MR-MEGA (Mägi et al., 2017) with 3 principal components of ancestry. We defined genome-wide significant loci by iteratively spanning the ± 500 kb region around the most significant variant and merging overlapping regions until no genome-wide significant variants were detected within ± 500 kb. The most significant variant in each locus is selected as the index variant. The nearest gene(s) to the index variant is used to name each locus. Cochran’s *Q*-test for heterogeneity has been conducted to identify loci with index variants that have different effect sizes across GWAS data sets, ancestry, or in males and females.

### PC projection

179,195 genetic variants have been genotyped/imputed in all biobanks, among which 168,899 are also in the 1000 Genomes (1000 Genomes Project Consortium et al., 2015) and HGDP (Cann et al., 2002). The weights corresponding to principal components for those markers were estimated based on the PCA analysis for the reference samples with known ancestry in 1000G and HGDP and shared among biobanks. Biobanks then generated PC loadings based on the pre-estimated weights of those markers.

### Variant annotation

We annotated genetic variants using ANNOVAR (Wang et al., 2010) for the nearest genes. To obtain a more complete annotation for putative loss-of-function variants, VEP (McLaren et al., 2016) with the LOFTEE plug (Karczewski et al., 2020) as implemented in Hail was used

### Heritability estimation and genetic correlation

We conducted LD score regression analyses using LDSC (Bulik-Sullivan et al., 2015) to estimate narrow-sense heritability based on the GWAS summary statistics for individual biobanks and to estimate the genetic correlation coefficients between each biobank and the all other biobanks together (LOBO). As most of the samples in the LOBO meta-analysis are of European ancestries, for biobanks with samples of non-EUR ancestries, such as BBJ, we estimated the trans-ancestry genetic correlation estimation using Popcorn(Brown et al., 2016) based on the LD scores pre-estimated using UK Biobank samples.

### Prioritize functional genes

#### DEPICT

Data-driven Expression-Prioritized Integration for Complex Traits (DEPICT) (Pers et al., 2015) was applied to investigate the results from genome-wide association studies of 14 endpoints. DEPICT uses three analyses to predict the gene functions: 1) prioritize the most likely causal genes, 2) identify enriched gene sets, and 3) discover tissues/cell types with highly expressed genes at associated loci. Two p-value thresholds were used to define genome-wide significance 1×10^−5^ and 5×10^−8^, for input summary statistics. A reference panel from individuals of European ancestry in 1000 Genomes was used to calculate LD and further identify the tag SNP from GWAS results. A minimum of 10 index variants from GWAS results was set to perform analysis using DEPICT. Enrichment results for significant findings from DEPICT were defined by FDR < 0.05. Sensitivity analysis was conducted with GWAS summary statistics derived from the meta-analysis of biobank data sets with samples of European ancestries (not including Finns) using LD information from the 1000 Genomes European panel to compare our findings with DEPICT results using multi-ancestry GWAS summary statistics.

#### PoPs

Polygenic Priority Score (PoPS) is a gene prioritization method used in our study to identify potential causal genes (Weeks et al., 2020). PoPS integrates GWAS summary statistics with publicly available bulk and single-cell gene expression, biological pathway, and predicted protein-protein interaction data to comprehensively perform gene prioritization. PoPS first applied Multi-marker Analysis of GenoMic Annotation (MAGMA) (de Leeuw et al., 2015) to meta-analyze gene-level associations and create gene-gene correlation matrix. Gene-level associations were generated by meta-analyzing the variants across the same gene, using GWAS summary statistics and LD panel from 1000 Genomes European only dataset. Next, MAGMA integrated previously calculated gene-level associations and gene-gene correlation to perform enrichment analysis for gene features selection. Lastly, a PoPS score was calculated by fitting a joint model with all the selected features simultaneously. In our study, genes with a PoPS score in the top one percentile were considered as the prioritized genes.

#### Transcriptome-wide association studies (TWAS)

Prediction of gene expression: Using genotypes and gene expression from 296 European donors from GTEx ver. 8 (Consortium, 2020), we trained predictive expression models using Joint-Tissue Imputation (JTI)(Zhou et al., 2020) and Multi-Omic Strategies for TWAS (MOSTWAS)(Bhattacharya et al., 2021b). Due to small eQTL sample sizes of non-European patients in GTEx, we restricted TWAS to European populations. We used gene expression from multiple relevant tissues for the analysis. For asthma, gene expression in Lung was used and for POAG, gene expression in Brain Cortex was used. For VTE, gene expressions in five most relevant tissues were used: Artery Aorta, Artery Coronary, Artery Tibial, Heart Atrial Appendage, and Heart Left Ventricle. JTI borrows information across transcriptomes of different tissues, leveraging shared genetic regulation, to improve prediction performance in a tissue-dependent manner (Zhou et al., 2020). MOSTWAS prioritizes distal-SNPs to a gene of interest that are mediated by biomarkers local to the distal-SNPs; these prioritized distal-SNPs are incorporated in the final model. We only considered genes with positive SNP heritability at p-value < 0.05 and adjusted cross-validation (CV) R^2^ > 0.01 with p-value < 0.05; we considered the gene model from the method that showed larger CV R^2^ for TWAS.

Association testing and probabilistic fine-mapping: Using meta-analyzed GBMI GWAS summary statistics from European-ancestry subjects, we detected gene-trait associations through the weighted burden test and 1000 Genomes Project CEU population as an LD reference (1000 Genomes Project Consortium et al., 2015; Gusev et al., 2016). We defined a transcriptome-wide significance using a Bonferroni correction across 20,000 tests (p-value < 2.5 × 10^−6^) (Gusev et al., 2016, 2018; Mancuso et al., 2017). As complex correlations between predicted expression levels at a given region can yield multiple associated genes in TWAS, we used FOCUS, a probabilistic gene-level fine-mapping method, to define credible sets of genes that explain the expression-trait signal at a given locus(Mancuso et al., 2019). Here, we used the default non-informative priors implemented in FOCUS and estimated the posterior inclusion probability (PIP) and a 90% credible set of genes at a given locus.

#### Proteome-wide Mendelian randomization (PWMR)

We estimated the putative causal role of 1,310 proteins on eight diseases in the NFE samples using proteome-wide association study (PWMR) and sensitivity analyses. For the exposure of the analysis, 5,418 conditional independent pQTLs of 1,310 proteins in European samples from ARIC (Zhang et al., 2021) were selected as genetic predictors. For outcomes, eight of the 14 diseases from GBMI were selected since they had full GWAS summary statistics in both European and African ancestries and had relatively good sample size (> 100 cases). The eight disease outcomes included idiopathic pulmonary fibrosis (IPF), primary open-angle glaucoma (POAG), heart failure (HF), venous thromboembolism (VTE), stroke, gout, chronic obstructive pulmonary disease (COPD) and asthma in European and African ancestries. For the discovery PWMR analysis, we applied a generalised inverse variance weighted approach (Burgess et al., 2017) that takes into account the correlation between genetic predictors. To increase the possibility of identifying true causal links between proteins and diseases, we applied five sensitivity analyses. First, we applied generalised MR-Egger regression to estimate the influence of horizontal pleiotropy (Burgess et al., 2017). For PWMR association with a p-value of the gEgger intercept term lower than 0.05, we considered these associations as influenced by horizontal pleiotropy and excluded them from the top finding list. Second, we applied Cochrane’s Q test for gIVW results and Rocker’s Q test for gEgger results to estimate the potential heterogeneity of PWMR estimates (Bowden et al., 2017; Greco M et al., 2015). Third, we applied three types of genetic colocalization analyses to distinguish causality from confounding by LD. The conventional colocalization, pairwise conditional and colocalization (PWCoCo) and LD check (Giambartolomei et al., 2014; Zheng et al., 2020). Fourth, to control for potential aptamer binding artificial effect of pQTLs, we listed all PWMR associations using pQTLs from the coding regions and flagged these associations with caution. Fifth, to estimate the influence of potential reverse causality, we applied MR-Steiger filtering (Hemani et al., 2017) and removed any PWMR associated with evidence of reverse causality from the top finding list. All the remaining PWMR associations with p-value < 0.001 were selected as candidate findings.

### Phenome-wide association test

For all 508 identified index variants (in known or novel loci), we carried out look-ups for their association with 1,283 human diseases curated based on phecodes mapped to ICD codes in the UK Biobank(Zhou et al., 2018). We reported associations with p-value < 5 × 10^−8^. The index variant was lifted over to GRCh37 for comparison with the UKBB results.

### Polygenic scores

The polygenic scores (PRS) were constructed using PRS-CS (Ge et al., 2019), which is based on the Bayesian framework. We used the auto model with default parameters implemented in the software to estimate the posterior mean SNP effects. The input for GWAS sample size was estimated as the total effective sample size. The LD matrix calculated using European individuals from 1000G Phase 3 (1KG) provided by PRS-CS was used here. Specifically, we used leave-one-biobank-out meta-analysis GBMI Asthma GWAS as the discovery GWAS and validated the PRS in 9 different biobanks, including: BBJ, BioVU, Lifelines, UKBB, CanPath, ESTBB, FinnGen, HUNT and MGI. To quantify the accuracy improvement attributable to GBMI, we also built PRS using public GWAS from (Demenais et al., 2017). The prediction performance of PRS was estimated using Nagelkerke’s *R*^2^ after regressing out the biobank-specific covariates with a logistic regression. It was further transformed to *R*^2^ on the liability scale(Lee et al., 2012), with biobank-specific case proportion used as the disease population prevalence. The corresponding 95% confidence intervals (CIs) were calculated using bootstrap with 1000 replicates.

## Data Availability

The all-biobank meta-analysis results and plots for the 14 endpoints (including both ancestry-specific and cross-ancestry meta-analyses and sex stratified meta-analyses) are available for downloading at https://www.globalbiobankmeta.org/resources and browsed at the PheWeb Browser http://results.globalbiobankmeta.org. Custom scripts used for quality control, meta-analysis and summary of results are available at https://github.com/globalbiobankmeta. The optimized trans-ancestry and single-ancestry polygenic score weights will be deposited within the PGS Catalog (https://www.pgscatalog.org/).

## Acknowledgments

The work of the contributing biobanks was supported by numerous grants from governmental and charitable bodies, and biobank specific acknowledgements are included in the **Supplementary Notes**. We would like to thank the organizing committee of the International Common Disease Alliance for intellectual contributions on the set up of the GBMI as a nascent activity to the larger effort. We would like to thank Daniel King from the Hail team and Sam Bryant from the Stanley Center Data Management team at the Broad Institute for helping with the Google bucket set up and data sharing, and Bethany Klunder from the University of Michigan Medical school for helping with the paper submission.

